# Magnetization Transfer MRI of intragastric milk digestion: a feasibility study in humans

**DOI:** 10.1101/2024.05.23.24307785

**Authors:** Morwarid Mayar, Camilla Terenzi, John P.M. van Duynhoven, Paul A.M. Smeets

**Affiliations:** Division of Human Nutrition and Health, Wageningen University & Research, Wageningen, The Netherlands; Laboratory of Biophysics, Wageningen University & Research, Wageningen, The Netherlands

## Abstract

Gastric milk protein (MP) coagulation has been extensively studied using *in vitro* and animal models. Yet, verifying these results in humans remains essential. Recently, we demonstrated that ^1^H Magnetization Transfer (MT) MRI can be used for the assessment of gastric MP coagulation *in vitro.* In the present follow-up study, we assessed the feasibility of using MT MRI for monitoring MP coagulation *in vivo* in humans. Twelve healthy adults underwent MRI scans before and after ingesting 300 g of low- or high-pasteurized skim milk (LPSM or HPSM, respectively). We assessed coagulation and gastric emptying (GE) dynamics by deriving the MT ratio (*MTR),* the total gastric content (TGC), semi-solid, and liquid volumes. The *MTR* values increased during digestion for both milk products, indicating a decrease in protein molecular mobility and a concomitant increase in the degree of coagulation. Prolonged heating of milk did not affect the *MTR* (p = 0.15), but resulted in higher TGC volumes (MD 40.3 mL, 95% CI [25.5-55.1], p = 0.044) with a trend towards greater semi-solid (p = 0.056) or liquid (p = 0.065) volumes. These findings suggest that prolonged heating of milk leads to slower GE. We thus showed that by combining MT with conventional anatomical MRI both MP coagulation and GE dynamics, and thereby the impact of heating on gastric milk digestion, can be assessed *in vivo* in humans. Our work underpins the feasibility of using MRI as a non-invasive imaging tool for studying the effect of food processing and composition on gastric digestion.

## 1. Introduction

There is an increasing socio-economical demand in linking food with health. To aid the development of innovative food products and processing methods for optimal health benefits, one of the necessary requirements is to understand the behavior of foods in our digestive system (Bornhorst & Paul, 2014). To this purpose, various static and (semi)-dynamic *in vitro* digestion models (Dupont & Mackie, 2015) have been developed, which simulate digestion in the human gastro-intestinal (GI) tract. Such models have found widespread use because they are simple to use, well controlled and do not pose ethical concerns. However, even sophisticated dynamic *in vitro* models do not fully capture the complexity of *in vivo* digestion, which includes neural and hormonal regulation as well as feedback controls. Therefore, *in vivo* data from humans is needed to gain a better understanding of food digestion, and to verify and inform *in vitro* digestion models. This requires the development of non-invasive methods that can be used to study food digestion both *in vitro* and *in vivo* in humans.

We have previously demonstrated that Magnetization Transfer (MT) ^1^H MRI is a powerful imaging approach for assessing structural changes that occur during static (Mayar et al., 2022, 2023) and semi-dynamic (Mayar et al., 2024) *in vitro* gastric digestion using cow’s milk, a widely consumed source of high-quality proteins, as a test case.

Cow’s milk contains approximately 3.5 w/v% protein, with caseins and whey proteins (WPs) constituting 80% and 20%, respectively (Bhat et al., 2016). Gastric digestion of cow’s milk involves acid- and pepsin-induced aggregation of casein micelles, which results in the formation of a semi-solid coagulum (Huppertz & Chia, 2021). The coagulation process results in reduced macromolecular mobility as liquid milk undergoes a transition into a semi-solid mass. In our prior *in vitro* work we demonstrated that this transition can be monitored using the ^1^H MT ratio (*MTR*) as an MRI marker of the coagulum consistency, and that differences in the coagulation behavior resulting from heat treatment can be assessed (Mayar et al., 2024).

Knowledge of the effect of heat treatment on gastric milk protein (MP) digestion is crucial because it can potentially be used to tune the gastric coagulation properties and the kinetics of the subsequent breakdown and absorption in the intestines. Therefore, gastric MP coagulation has been studied extensively with *in vitro* models (Li et al., 2022a; Mulet-Cabero et al., 2019; Ye et al., 2016a) and *in vivo* in animals (Ahlborn et al., 2023; Ye, Liu, et al., 2019). These studies have shown that heat treatment results in a looser and softer coagulum consistency under gastric conditions, due to aggregation of WPs on the surface of casein micelles, resulting in reduced casein-casein interactions (Kethireddipalli & Hill, 2015). Moreover, a study in humans reported a higher dietary nitrogen (N) transfer into serum amino acids (AAs) and proteins for UHT compared to pasteurized milk (Lacroix et al., 2008). The authors suggested that this may have been caused by the looser and softer coagulum structure and faster gastric emptying (GE) of the UHT milk. However, recently, Milan et al. (2024) showed that GE measured by MRI was slower for UHT compared to pasteurized milk. Nevertheless, gastric coagulum formation and degradation were not directly considered in their study, and to date there are no *in vivo* human studies that have assessed the effect of heat treatment on intragastric MP coagulation. As demonstrated by our *in vitro* results, ^1^H MT MRI holds great potential for bridging this research gap (Mayar et al., 2022, 2023). However, studying gastric digestion is more challenging *in vivo* than *in vitro,* due to motion caused by breathing and gastric motility, as well as to large biological variation. Therefore, the first aim of this study was to assess the performance of ^1^H MT MRI for assessing gastric MP coagulation in humans using low- and high-pasteurized skim milk (LPSM and HPSM, respectively) as test products. Since the consistency of the coagulum may also affect GE, the secondary objective was to compare GE dynamics of LPSM and HPSM in terms of changes in total, semi-solid and liquid gastric content volume using *T*-weighted anatomical MRI images.

## 2. Materials and methods

### 2.1. Study design

The study was a single-blind randomized crossover trial in which healthy, normal-weight adults underwent gastric MRI scans at baseline and after consumption of two differently heated skim milk products. The primary outcome was the ^1^H *MTR* of the gastric content over time. Secondary outcomes were the total gastric content (TGC), the semi-solid and liquid volume fractions over time. Other outcomes were subjective ratings of appetite (hunger, fullness, thirst, desire to eat) and well-being (nausea and bloating). The study procedures were approved by an accredited Medical Ethical Committee (METC Oost-Nederland) and were in accordance with the Helsinki declaration of 1975 as revised in 2013. The trial was registered with clinicaltrials.gov under number NCT05854407. All participants provided written informed consent.

### 2.2. Participants

The study was conducted between June 2023 and September 2023 with 12 healthy (self-reported) adults (n = 6 males and n = 6 females, age 24 ± 4 years, body mass index (BMI) 22±2 kg/m^2^; Fig. S1 in the Supplementary Information (SI)). The sample size estimation can be found in the SI. Participants were recruited in the Ede and Wageningen area in The Netherlands via digital advertisements (email and Wageningen University website). The inclusion criteria were: age between 18 and 45 years; BMI between 18.5 and 25 kg/m^2^; self-reported good general health. The main exclusion criteria were: lactose and milk protein intolerance or allergy; use of medication that may alter the normal functioning of the digestive tract; having a gastric disorder or regular gastric complaints (≥ 1 per week) and having a contra-indication for MRI including, but not limited to, pacemakers and defibrillators, ferromagnetic implants, or claustrophobia. Potential participants were informed about the details of the study via an online information meeting, followed by a screening session which involved tasting of the milk products, getting accustomed to undergoing MRI scans in a dummy MRI scanner, practicing drinking milk in a supine position and practicing the breath holds required for the MRI scans. Volunteers that were still interested in participation were asked to sign the informed consent form and fill in the screening questionnaire.

### 2.3. Milk products

Commercial pasteurized skim milk (typically heated at 72 °C for 15 s) was purchased from a grocery store, and is referred to as Low Pasteurized Skim Milk (LPSM). High Pasteurized Skim Milk (HPSM) was prepared from LPSM by heating the latter in a water bath for 30 min after reaching a temperature of 80 °C. The WP denaturation level in the milk products was measured using SDS-PAGE (data not shown), and was 3% and 90% for LPSM and HPSM, respectively. The milk products were purchased or prepared within one week prior to the test session and stored at 4 °C. The milk was kept at room temperature for 30 min before the start of the test session. LPSM from the same brand was used throughout the study.

### 2.4. Study procedures

The participants visited the Hospital Gelderse Vallei in Ede, the Netherlands, twice in the morning between 7.30 and 9.30 am, in fasted state. There was a minimum of 1 and a maximum of 4 weeks between the two visits. Eating and drinking was allowed until 8:00 pm the night before, and drinking water was allowed up to 1 hour before the start of the test session. A schematic overview of the study session is shown in Fig. 1. Upon arrival, participants completed an MRI screening form. After that, a baseline abdominal MRI scan was conducted and the participants provided baseline verbal ratings of their appetite and well-being. Subsequently, participants drank 300 g (291 mL) of either LPSM or HPSM through a tube (similar to drinking from a straw), while lying in a supine position on the scanner bed. The mean ingestion time, with its standard deviation (SD), was 1.2 ± 1.1 min. MRI scans of the abdomen were performed at t = 5 min and subsequently at intervals of 15 mins up until 95 min after the start of ingestion. The participants were asked to verbally rate their appetite and well-being, on a scale between 0 and 100, after the MRI scanning at each digestion time. They remained in a supine position for the full duration of the MRI scanning. After this, they exited the scanner and were offered a takeaway breakfast.

**Figure 1.**
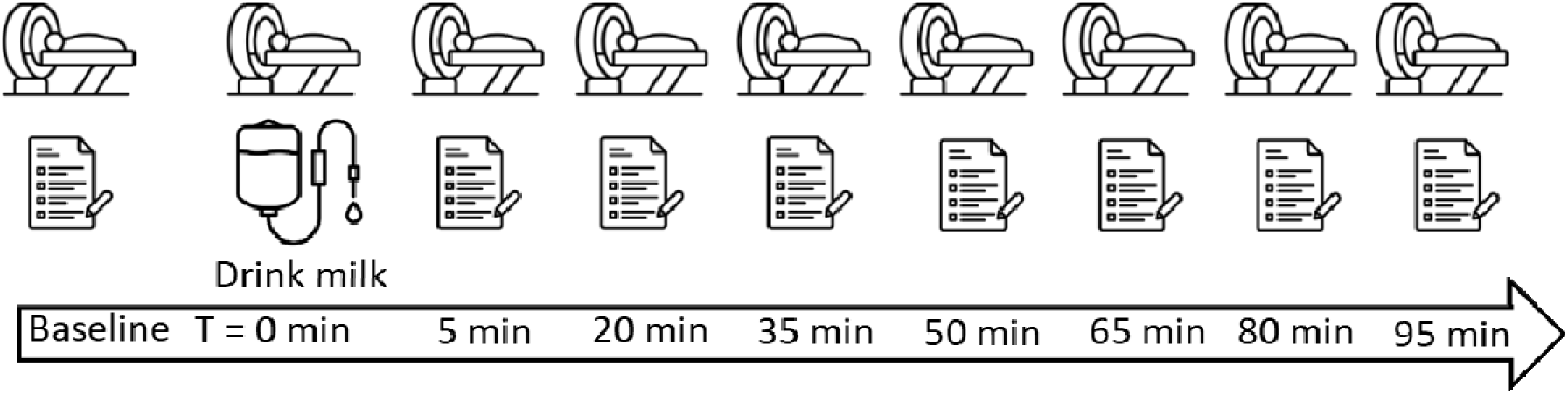
Schematic overview of a test day consisting of MRI baseline measurements, drinking 300 g (291 mL) of milk in a supine position through a tube (similar to drinking from a straw), followed by MRI measurements subjective ratings of appetite and well-being after 5 min from milk ingestion, repeated every 15 min up to 95 min.

### 2.5. MRI scans

Participants were scanned using a 3T MRI scanner with the dStream torso coil (Philips Ingenia Elition X, Philips Medical Systems, the Netherlands). All scans were performed during breath-holds. Participants were asked to hold their breath on expiration to minimize motion. The breath hold duration was at most 21 s. At each timepoint, Saturation Transfer (ST) and -weighted measurements were conducted. The spatial variation in the RF field () was determined by measuring a flip angle map, using the dual refocusing echo acquisition mode (DREAM) sequence (Nehrke & Börnert, 2012). The flip angles were divided by the nominal flip angle to map the relative irradiation amplitude (). The H MT MRI measurements were conducted using a saturation RF pulse combined with a Rapid Acquisition with Refocusing Echoes (RARE) sequence. A reference unsaturated MRI image (and a corresponding saturated image (were acquired at frequency offsets () of 450 ppm and 10 ppm, respectively in order to obtain the. In addition, MRI images were acquired at = 2.7 and -2.7 ppm to obtain the maps and Water Saturation Shift Referencing (WASSR) spectra were acquired to construct a -map (data not shown in this paper). The saturation pulse consisted of a train of pulses with a total duration (of 1 s and a saturation pulse amplitude (of 3 µT. Three sagittal slices with a field-of-view (FOV) of 400 mm x 352 mm, in-plane resolution of 1 mm x 1 mm, slice thickness of 4 mm, slice gap of 2 mm were acquired. SINC pulses were used for excitation and refocusing. The excitation flip angle was 90° and the first refocusing flip angle was 180° followed by 110° pulses. A repetition time () of 6.6 s, an effective echo time (*TE_eff_*) of 80 ms, and a turbo factor (*TF*) of 82 were used, resulting in a total acquisition time of 79.5 s. To assess TGC, semi-solid and liquid volumes, *T_2_*-weighted anatomical images were acquired using a RARE sequence with 28 axial slices, FOV of 400 mm x 400 mm, in-plane resolution of 0.625 mm x 0.625 mm, slice thickness of 4 mm and inter-slice gap of 1.4 mm. The excitation and refocusing RF pulse angles were the same as for the MT scans. The adopted MRI acquisition parameters, namely *TR* of 755 ms, *T*_eff_ of 80 ms, and *TF* of 65, resulted in a total acquisition time of 21 s. Shimming and pulse calibrations were conducted before the MT and *T*-weighted anatomical scans at each measured digestion timepoint. Shimming was performed on the stomach area using pencil beam volume with high-order shims.

### 2.6. MRI data processing and analysis

The TGC was manually delineated in the *S*_o_ and *S*_sat_ MT MRI images for each slice at each timepoint using the Medical Image Processing And Visualization (MIPAV) software 11.0.3. (National Institutes of Health, Bethesda, MD, USA) and stored as a binary mask. All further image processing and calculations were conducted in MATLAB 2019b (MathWorks, Massachusetts, USA). The masks were used to extract the gastric content. Rigid (translation and rotation) and non-rigid (deformation) motion of the stomach were corrected for by using the robust principal component analysis (RPCA) approach (Bie et al., 2019), combined with affine image registration. The image correction was performed after applying the TGC mask on the images because this provided better spatial alignment of the gastric content between the reference *S*_o_ and *S_sat_* MRI images compared to performing the correction on the whole FOV. In short, RPCA was used to decompose the *S*_0_ and *S*_sat_ (including at Δ ± 2.7 ppm) images into a low-rank and sparse component of the image to separate the contrast from the motion. The motion-free low-rank component was recomposed into the *S*_0_ and *S_sat_* images, and the average of these images was calculated to obtain a motion-free reference image. The reference image was used for re-alignment of the original motion-corrupted images (Fig. S2 in SI). The re-aligned gastric content images were used to obtain masks of the low- and high-intensity image voxels, attributed to the semi-solid and liquid components, respectively. This segmentation was achieved via automatic intensity-thresholding using the multi-thresh function (Otsu’s method) with two levels in MATLAB, similar to the approach used on in vitro images of milk digestion (Mayar et al., 2024). Otsu’s method finds the optimal threshold value that groups the voxels into separate classes for which the within-class variance is minimised. Voxels with intensities below or above the second threshold level, which represented 40-50% of the maximum image intensity, were respectively categorized as low- or high-intensity voxels, and attributed to semi-solid or liquid fractions (Fig. S3 in SI). The low-intensity voxels were saved as a binary mask and applied to the re-aligned MT images to calculate the *MTR* of the semi-solid content for each slice using Eq. 1, from which we can see that the *MTR* ranges between 0 and 1.

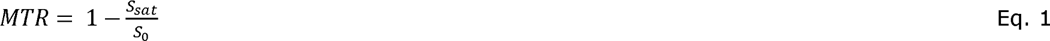

The experimental *MTR* values of the semi-solid content were corrected by omitting values that were <0 and ≥1, which are likely caused by gastric content mixing. The corrected data for each slice was used to calculate the mean and SD of the *MTR* over all the low-intensity (semi-solid) voxels for each time point and participant. The *MTR* in the liquid fraction is negligible because of the high ^1^H mobility. The SD of the *MTR* map was calculated as a measure of the spatial variation in the semi-solid gastric content *MTR* values.

Gastric content masks of the *T_2_*-weighted scans in the axial plane were obtained in the same way as described for the MT scans. These masks were used to calculate the TGC volume by multiplying the total number of voxels by the individual voxel volume, taking into account the slice thickness and the inter-slice gap. GE was defined as the decrease in TGC volume over time. The semi-solid and liquid masks were obtained in the same manner as for the MT data and were subsequently used to calculate the corresponding volumes.

### 2.7. Statistical analysis

Data are reported in the form mean ± SD unless stated otherwise. All statistical analysis were performed using RStudio 4.3.1 (PBC, Boston, MA). The threshold for statistical significance was set at p = 0.05. Normality of the data was checked with quantile-quantile plots of the residuals. A Sidak-adjusted Linear Mixed Model (LMM) was used to test for effects of time, treatment and a treatment-by-time interaction on the *MTR* of the gastric content. Time and treatment (milk products) were added as fixed effects and participants were included as a random effect. Outliers in the MT data were identified and removed using the 3*IQR (interquartile range) criterion method (Fig. S4 in SI). A LMM was also used to test the effect of time, treatment and treatment-by-time interaction on the TGC, semi-solid and liquid volumes with the inclusion of baseline TGC volumes as a covariate. For the appetite and well-being ratings, baseline ratings were included as a covariate. In the case of a significant treatment effect, Tukey’s HSD post-hoc t-tests were used to compare individual time points. As an exploratory analysis, the effect of sex on the outcomes was tested using a LMM with time and sex as fixed factors, baseline GC volume as a covariate and the participants as a random factor. The two treatments were grouped per sex. The analysis was conducted with and without the inclusion of body size, approximated as weight times height of the participant, as a covariate. Paired t-tests were used to compare LPSM and HPSM with the following measures: the incremental area under the curve (AUC) of the *MTR*, TGC, semi-solid and liquid volume, as well as the *MTR*_max_ and the change in the *MTR* between t = 5 min and t = 95 min (*LlMTR*).

## 3. Results

### 3.1. TGC, semi-solid and liquid volume vs. digestion time

The ^1^H MRI *T*_2_-weighted images were used to quantify the volume of the TGC and semi-solid or liquid fractions over time (Fig. 2). At baseline, there was only gastric juice present and, at t = 12 min, the milk was still in the liquid state. As digestion progressed, low-intensity voxels at the center surrounded by high-intensity voxels at the stomach wall were observed, respectively, corresponding to the semi-solid coagulum or gastric fluid, and indicated in the insets in red or blue. The baseline GC volume was 31 ± 18 mL and 35 ± 12 mL for LPSM and HPSM treatments, respectively (p = 0.69). After milk ingestion, the TGC volume correspondingly increased to 267 ± 44 mL and 288 ± 40 mL for LPSM and HPSM, respectively, followed by a decrease over time (Fig. 3a). There was a significant effect of treatment on the TGC volume, with lower volumes for LPSM (p = 0.044, MD = 40.3 mL 95% CI [25.5 – 55.1]). This effect was significant at most timepoints (t = 27 and 57 – 102 min, all p<0.05). There was no treatment-by-time interaction effect (p = 0.49). The semi-solid volume (Fig. 3b) decreased linearly with time for both LPSM and HPSM. The treatment effect tended to be significant (p = 0.057) with lower volumes for LPSM (MD = 31.8 mL 95% CI [19.9-43.8]), and the treatment-by-time interaction effect was not significant (p = 0.26). The liquid volume (Fig. 3c) for LPSM decreased with a similar trend as the TGC volume, with lower volumes compared to HPSM from 57 min onwards. The liquid volume for HPSM showed little variation over time. There was no main effect of treatment on the liquid volume (p = 0.68), but the treatment-by-time effect tended to be significant with higher liquid volumes for HPSM at t ≥57 min (p = 0.065, MD = 65.8 mL 95% CI [43.4 – 88.2]).

**Figure 2.**
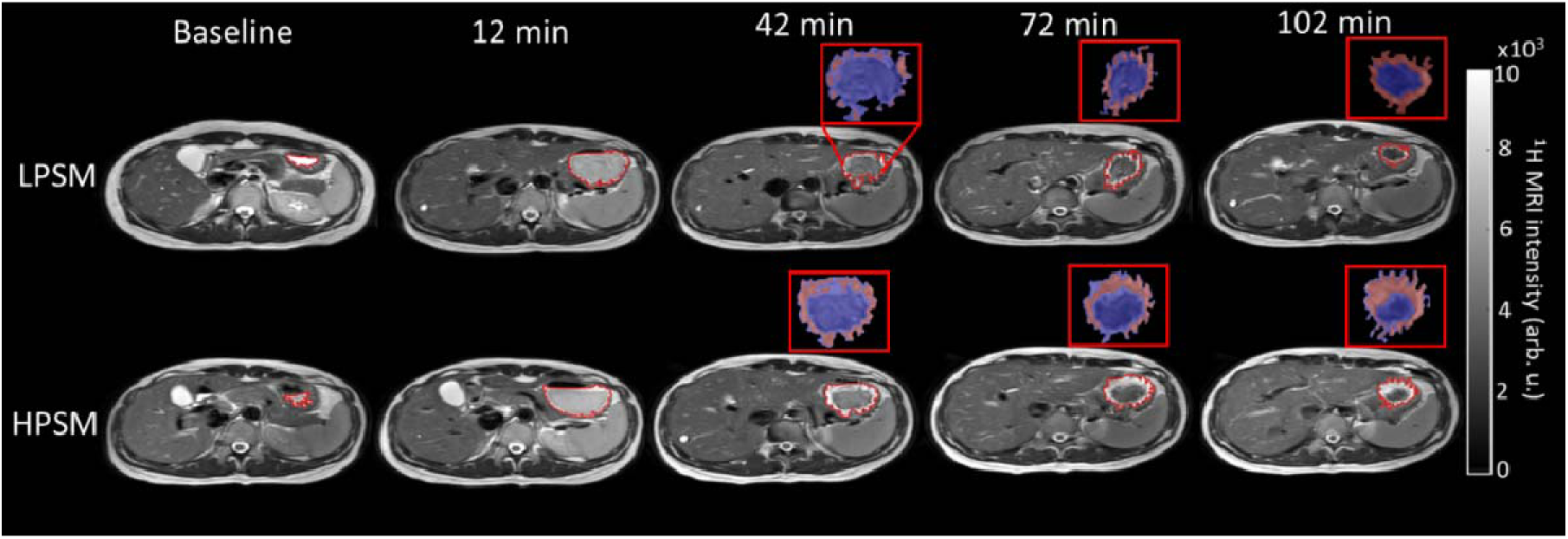
Example of transversal ^1^H MRI-weighted images at baseline and during digestion, showing the emptying of the stomach and the changes in liquid (red) and semi-solid (blue) fractions as shown in the insets on the top right.

**Figure 3.**
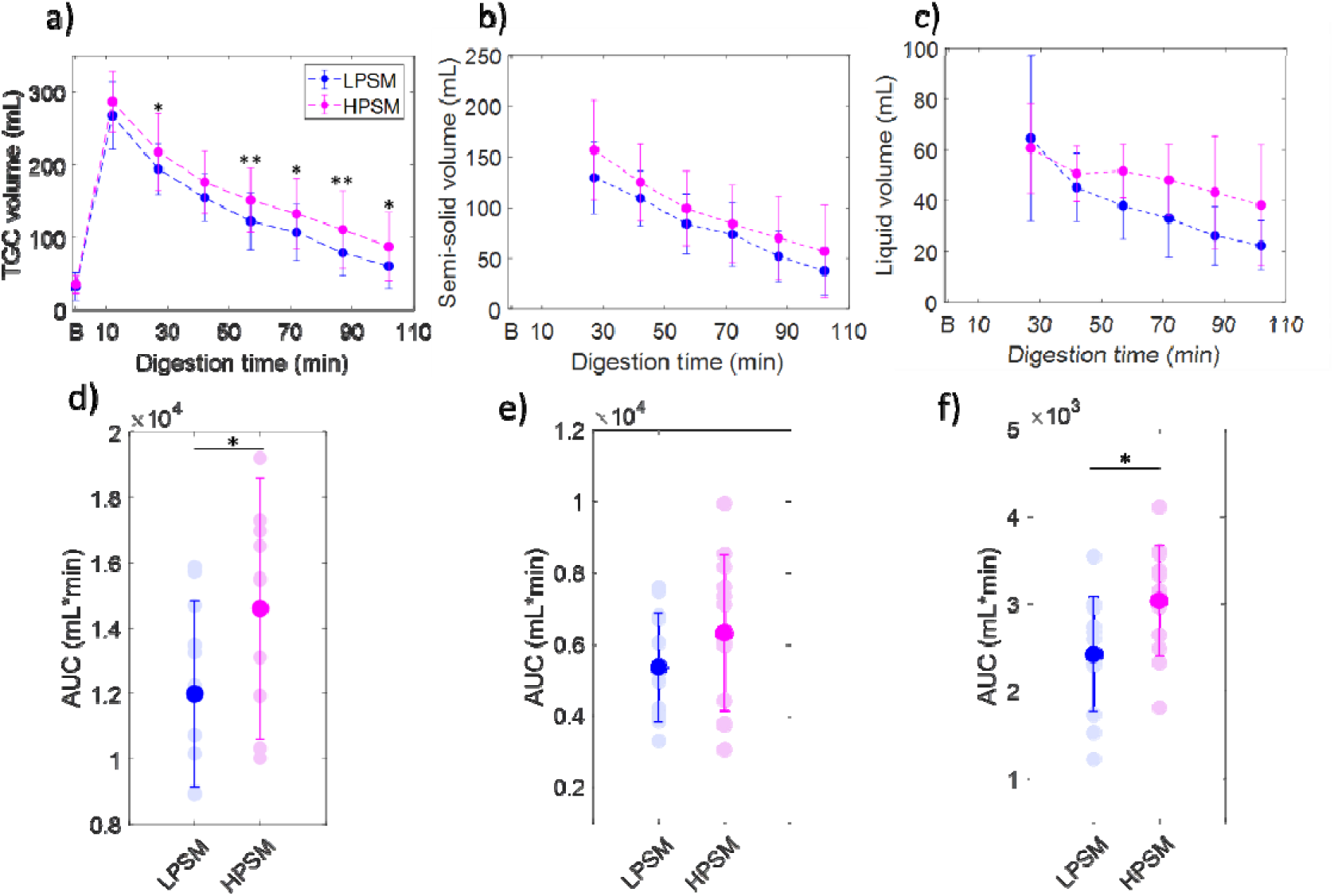
(a) TGC, (b) semi-solid and (c) liquid volumes vs. digestion time for LPSM and HPSM estimated from the -weighted MRI images, along with the respective AUC of the (d) TGC, (e) semi-solid and (f) liquid volumes. Symbols ** and * stand for p<0.01 and p<0.05, respectively. Data are plotted as mean ± SD over all participants. In d-f, individual data for each participant are also shown.

The AUC of the TGC (Fig. 3d) and liquid volume (Fig. 3f) were lower for LPSM compared to HPSM (p = 0.021 and p = 0.017). There was a trend towards a lower semi-solid volume for LPSM (p = 0.078) (Fig. 3e).

#### 5.3.2. ^1^H MT MRI during gastric digestion

The sagittal-weighted images of the abdomen (top), and the corresponding maps overlayed on the -weighted images (middle), are shown in Fig. 4. At baseline and t = 5 min, the gastric content appeared homogeneous within each-weighted image (Fig. 4a). As digestion progressed, the size of the stomach decreased due to GE and the semi-solid coagulum and liquid fractions could be observed. The maps (Fig. 4b) show that the at baseline was low, which is expected for the fasted state, where the gastric content is mainly composed of gastric juice. The at t = 5 min after the start of ingestion was higher than that at baseline due to the presence of the milk proteins, and subsequently increased with digestion for both LPSM and HPSM. The histograms of the values for each map (Fig 4c) were characterized by a narrow unimodal distribution at t = 5 min, while such distribution broadened and shifted towards higher values at longer digestion times.

**Figure 4.**
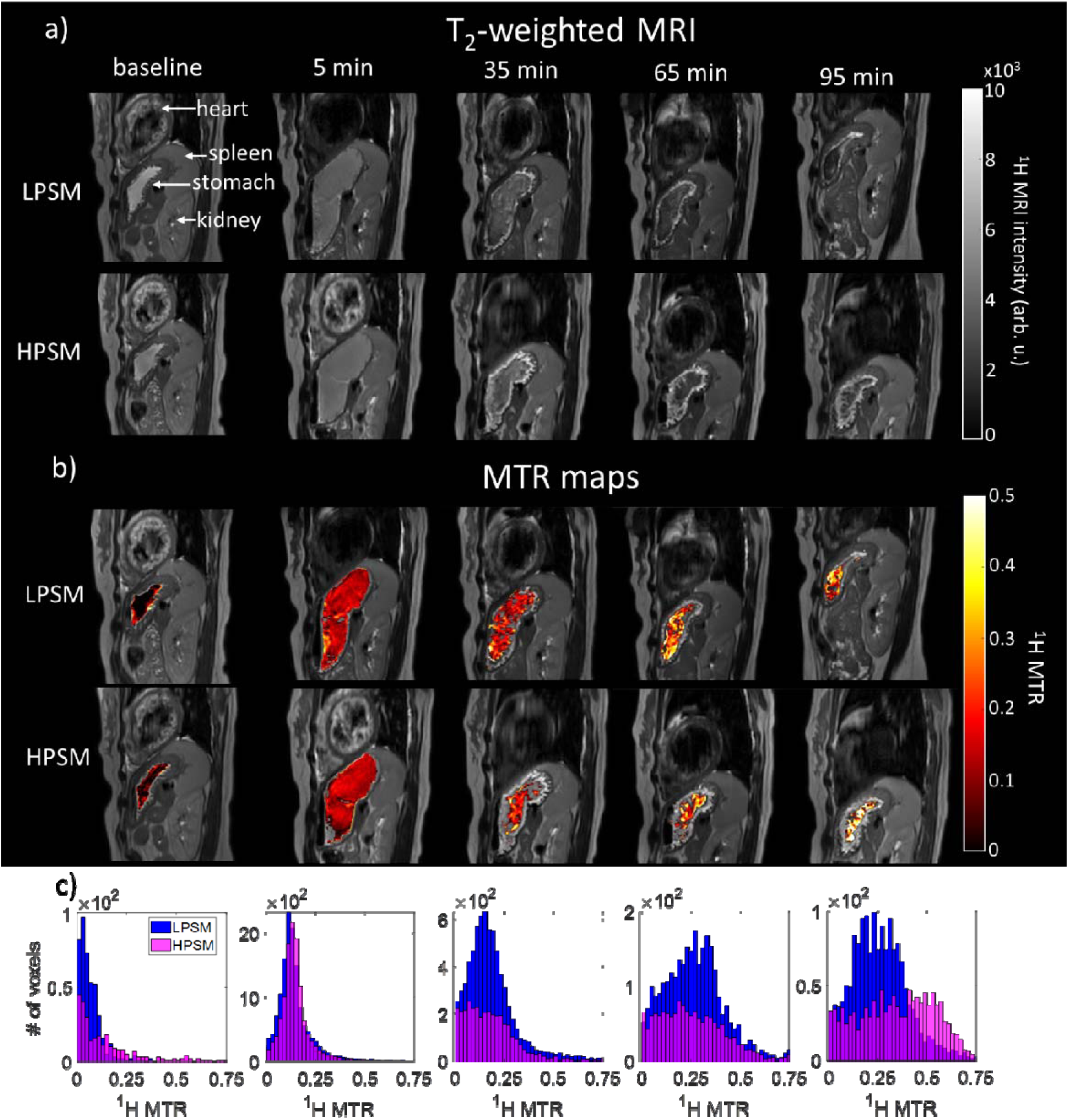
(a) -weighted sagittal MRI images of the abdomen at baseline, t = 5 min after ingestion of 300 g of LPSM or HPSM and at 30 min intervals until t = 95 min. The ^1^H MRI intensities are scaled to the maximum intensity for each individual image. Corresponding (b) color-coded maps and (c) histograms in which the y-axis is scaled to the gastric content volume for each time point. Data are from one randomly selected participant.

The mean *MTR* increased over time for both LPSM and HPSM (p<0.001, Fig. 5a). The MD between LPSM and HPSM was 16% (95% CI [10-21]). The *MTR* values of HPSM appeared higher than those of LPSM from 35 to 65 min. However, no significant treatment or treatment-by-time interaction effects were found (p = 0.15 and p = 0.58, respectively). The spatial variation of the *MTR* values also increased with time, which may indicate an increase in the heterogeneity of the semi-solid gastric content with digestion (Fig. 5b), in agreement with the observations from the histograms in Fig. 4. In contrast to the *MTR* values, the average *rB_1_* and its spatial variation over the voxels used for calculating the *MTR* maps, decreased with the digestion time from 100 ± 15% at t = 5 min to 87 ± 9% at t = 95 min (Fig. S5 of SI). As demonstrated in Fig. 5c-e and Fig. S6 in SI, the inter-individual variation in the *MTR* values was large, and there was no significant difference in the Δ*MTR* (p = 0.29), AUC *MTR* (p = 0.48) and *MTR*_max_ (p = 0.15) between LPSM and HPSM. The baseline *MTR* value tended to be lower (p = 0.057) for the LPSM (*MTR* = 0.08 ± 0.02) compared to HPSM (*MTR* = 0.10 ± 0.04) treatment. However, there was no correlation between the baseline *MTR* values and the AUC *MTR* or *MTR*_max_ (all r <0.35 and p >0.3; see Table S1).

**Figure 5.**
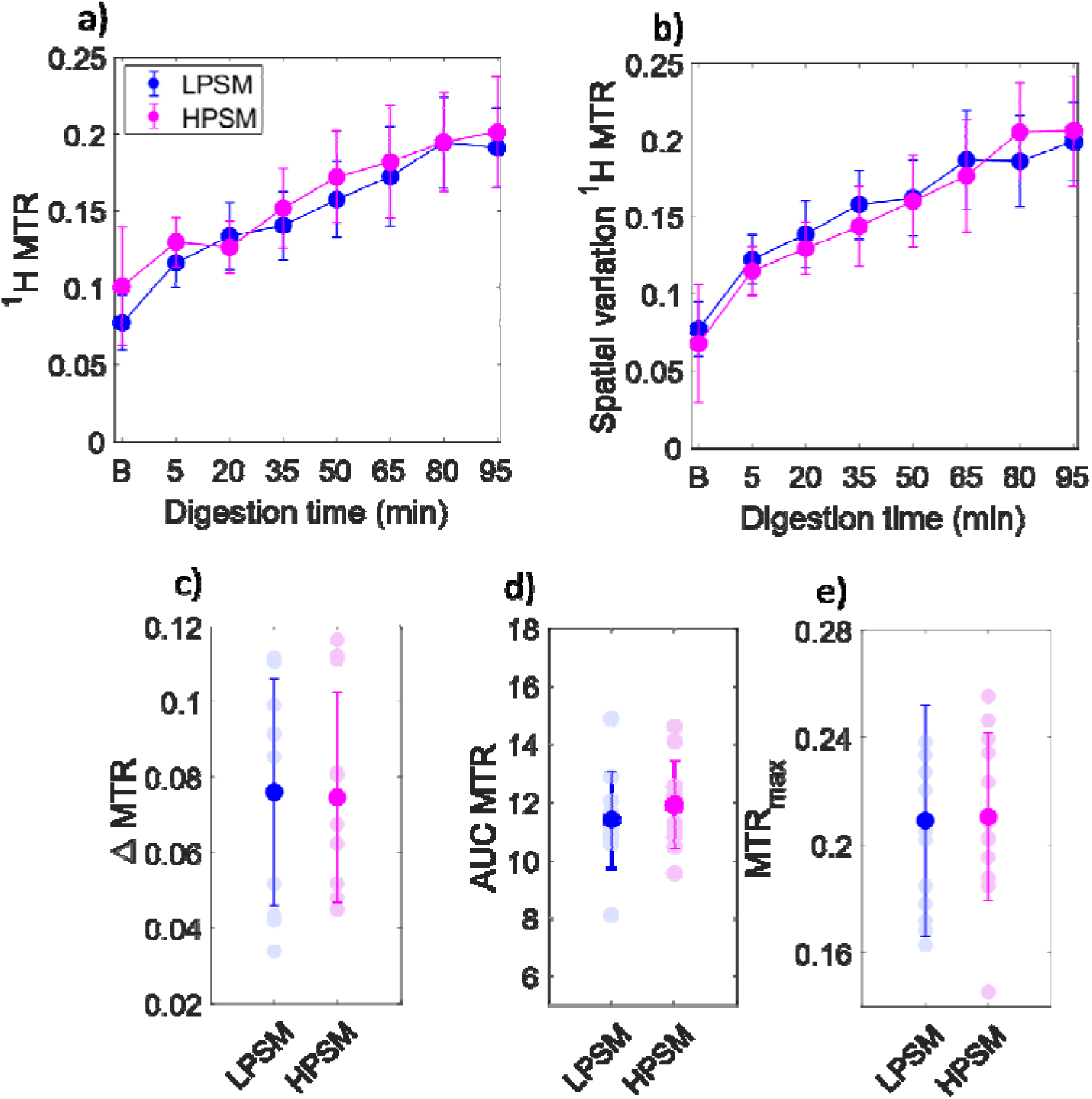
(a) Mean and (b) spatial variation, determined by the SD of the H values of the semi-solid voxels over time (c) The change in from 5 to 95 min after milk ingestion (Δ=(t=95)- (t=5)), (d) the AUC and (e) the. The data in (a) and (b) are plotted as the mean ± SD over all participants, and in c-e as mean ± SD for both treatments. In c-e individual data for each participant are also shown.

### 3.3. Exploratory analysis on the effect of sex on the outcomes

Exploratory analysis showed that the, TGC volume and liquid volume over time were higher for females compared to males (p<0.001, p = 0.025 and p= 0.026, respectively, Fig. S7 in SI). There was no difference in the semi-solid volume between males and females (p =0.12). The male and female participants had an average body size, estimated as weight times the height, of (1.4 ± 1.7)·10^4^ and (1.1 ± 0.9)·10^4^ kg*cm, respectively. After accounting for the effect of body size, the effect of sex on the *MTR*, TGC volume and liquid volume was no longer significant (p=0.46, 0.28 and 0.74, respectively).

### 3.4. Appetite and well-being ratings

The appetite and well-being ratings over time are reported in Fig. S8 in SI. Participants reported a higher level of hunger and appetite (p < 0.001) and a lower level of fullness (P<0.05) for HPSM compared to LPSM. For hunger and appetite, the effect was driven by most time points (p <0.05), whereas for fullness, the effect was driven by time points 42 min and 57 min (p = 0.02 and p = 0.048, respectively). There was no treatment effect on thirst, bloating or nausea (p = 0.31, p = 0.29 and p = 0.90, respectively). There was a significant treatment-by-time effect with higher ratings of nausea at t≥57 min for HPSM compared to LPSM (p = 0.04). However, this effect was mainly driven by 2 participants who reported 20 points higher nausea scores for HPSM compared to LPSM, whereas the other participants reported nausea scores of around 0 throughout the digestion for both milk products. Therefore, no firm conclusions can be drawn from this.

## 4. Discussion

The objective of this study was to assess the feasibility of using ^1^H MT MRI for monitoring gastric milk protein coagulation *in vivo* in humans using LPSM and HPSM as test-products. The TGC, semi-solid and liquid volumes were compared between the two products to investigate the effect of heat treatment on the GE dynamics.

### 4.1. GE dynamics

We found that the TGC volume remained higher for a longer duration for HPSM compared to LPSM. This suggest a slower GE with more extensive heat treatment. Additionally, the semi-solid and liquid volumes vs. digestion time appeared to be higher for HPSM compared to LPSM, although these differences were not statistically significant.

These findings contradict our initial expectation, as previous *in vitro* (Li et al., 2022; Mulet-Cabero et al., 2019) and *in vivo* animal (Ahlborn et al., 2023; Ye et al., 2019) studies have demonstrated that heat treatment typically results in a coagulum with a looser and softer consistency during gastric digestion, which has been suggested to potentially lead to a faster GE. Ahlborn et al. (2023) showed that, for a 500 mL load of milk, the TGC, coagulum, total protein and coagulated protein emptied faster for UHT compared to pasteurized milk during gastric digestion in pigs. These differences were attributed to the weaker and more open coagulum structure observed for UHT milk. In contrast, Ye et al. (2019) reported that the wet and dry weight of the coagulum were higher for UHT compared to pasteurized milk in rats at 30 and 120 min after milk ingestion, but slightly lower at 240 min. This suggests that the volume and moisture content of the coagulum from UHT milk is higher in the first 120 min of digestion, aligning with the higher semi-solid and liquid volume found for HPSM in the present study. The higher liquid volume for HPSM may indicate a higher moisture content which, in turn, could be attributed to a coagulum microstructure with more abundant and larger voids (Ye et al., 2016). Barbé et al., (2013) found that the mean retention time of chromium in the stomach in pigs was longer for heated (10 min at 90 °C) compared to unheated rehydrated ultra-low heat skim milk powder, which may suggest a slower emptying of milk upon heat treatment. Overall, *in vitro* and *in vivo* studies in animals have shown that high temperature or prolonged heat treatment result in a looser and softer milk protein coagulum during gastric digestion, but there is no consistent information on how this affects GE.

A recent *in vivo* study in humans showed that GE was slower for UHT compared to pasteurized milk, in line with the outcome of our study (Milan et al., 2024). They also showed that the concentration of essential AAs in the blood was higher for UHT compared to pasteurized milk. While other *in vivo* studies in humans (Horstman et al., 2021; Lacroix et al., 2008) did not report a difference in AA concentrations in blood following the consumption of UHT and pasteurized milk, Lacroix et al. (2008) did report an enhanced transfer of dietary nitrogen into serum AA and protein pools for UHT compared to pasteurized milk. They proposed that a softer coagulum and more rapid GE could explain the observed difference. However, as shown in the present study and by Milan et al. (2024), more extensive heat treatment resulted in a slower GE. This suggests that the differences in plasma AA concentrations observed after consumption of differently heated milk products is more likely caused by the higher gastro-intestinal hydrolysis level of denatured WPs from UHT milk than by differences in GE and coagulation dynamics.

### 4.2. Interpretation of MT MRI data

Following ingestion, the *MTR* was initially low but, as digestion progressed, a notable increase in the *MTR* was observed for both LPSM and HPSM (Fig. 5). Right after ingestion, the milk is in a liquid state and, consequently, the macromolecular mobility is still high, resulting in a low *MTR* (Henkelman et al., 2001; van Zijl et al., 2018). As digestion proceeds, a transition from a liquid to semi-solid state occurs, primarily driven by the aggregation of caseins induced by pepsin and stomach acid (Huppertz & Chia, 2021). This transition to a semi-solid state can cause a decrease in the macromolecular mobility, leading to stronger inter- and intra-molecular dipolar interactions, which are the drivers of the ^1^H MT effect in MRI (Henkelman et al., 2001; van Zijl et al., 2018; Zhou et al., 2023). In our preceding study (Mayar et al., 2024), a similar increase in the *MTR* was observed for LPSM during semi-dynamic *in vitro* gastric digestion. The *in vitro MTR* data was compared with visual and rheological assessment of the coagula, which confirmed that the increase in *MTR* corresponded to an increase in the storage modulus and, hence, in the stiffness of the coagulum. The agreement between the *in vitro* and *in vivo* data for LPSM, alongside the significant *MTR* variation vs. digestion time, confirmed the suitability of MT MRI for monitoring gastric milk protein coagulation *in vivo*. However, we did not observe an effect of heat treatment on the *MTR* over time. Interestingly, this is in contrast with our previous *in vitro* study (Mayar et al., 2024) where, based on visual assessment, the coagulum of HPSM appeared more loose compared to that of LPSM. This difference was also reflected in the *MTR* values, which were lower for HPSM compared to LPSM beyond 50 min of gastric digestion.

The *MTR* and GE data altogether suggest that the effect of heat treatment on gastric coagulation may be different in humans compared to what has been observed in *in vitro* and *in vivo* animal models. Gastric coagulation of milk is highly affected by pH and pepsin activity, which can vary both between and within individuals (Fadda et al., 2022; Semple et al., 2001). Furthermore, gastric fluid volume in the fasted state also varies between and within individuals (Grimm et al., 2018; Roelofs et al., 2024), and has previously been shown to affect the formation of a fat layer during gastric milk digestion (Camps et al., 2021). Accordingly, the gastric fluid volume may also affect gastric milk coagulation. In this study, the baseline *MTR* values tended to be lower for the LPSM compared to the HPSM treatment, which could have affected the coagulation process and, hence, the *MTR*. However, there was no correlation between baseline *MTR* values and the AUC *MTR* or *MTR*_max_ values. Consequently, additional research with larger sample sizes is required to provide more insights into the coagulation process.

A recent study assessed gastric milk coagulation in women experiencing gastro-intestinal (GI) complaints and in a control group after milk consumption by calculating image texture metrics, namely homogeneity, contrast, busyness, and coarseness, for the *T_2_*-weighted anatomical images (van Eijnatten et al., 2023). The decrease in homogeneity, and increase in contrast and coarseness, were associated with the coagulation process. While such image texture metrics show promise for monitoring gastric milk coagulation, they need to be verified with MRI data of *in vitro* digestion samples, analogously to what we have done for the *MTR* parameter.

### 4.3. Challenges in acquisition and quantification

Conducting MT measurements *in vivo* in humans is challenging due to artifacts caused by breathing motion and gastric motility. To construct the *MTR* maps, the *S*_O_ and *S*_sat_ images were acquired separately in different breath holds. Although breath holding minimizes breathing-related motion artifacts within an image, it does not account for differences between the *S*_O_ and *S*_sat_ images. Differences in both the position and shape of the stomach were observed, with the latter being caused by gastric contractions. These differences were corrected using rigid and non-rigid image registration. However, in certain cases, gastric contractions and mixing resulted in differences in the content of individual voxels between the *S*_O_ and *S*_sat_ images. For example, some voxels contained low-intensity components in the *S*_O_ image, arising from semi-solid fractions, but high-intensity (liquid) components in the *S*_sat_ image, leading to negative *MTR* values. These values were excluded from the *MTR* maps and from the corresponding calculation of the mean *MTR* over the semi-solid voxels. In future applications it is preferable to work on reducing the acquisition time of the scans to enable acquisition of both the *S*_O_ and *S*_sat_ images in one breath hold. This could help to minimize the sensitivity to mixing effects, and to improve the accuracy of the *MTR* measurements.

In this work, the ^1^H MT MRI scans were acquired using a RARE sequence with a 90° RF excitation flip angle and *T_2_*-weighting, with an acquisition time of around 19 s per image. In humans the stomach typically contracts about three times per minute (Lu et al., 2022), which suggests that the *S*_O_ and *S*_sat_ scans may have been acquired during two different contractions. The acquisition can be accelerated by replacing the RARE sequence with a faster sequence, such as a Gradient Recalled Echo (GRE) sequence. However, faster image acquisition schemes often come at the cost of lower signal-to-noise ratios (SNR) and poorer contrast. This could make it challenging to accurately delineate the gastric content and to separate liquid and semi-solid components using intensity thresholding. Therefore, further optimization of the scan parameters is required to achieve faster measurements, while maintaining a high SNR and good contrast. Faster measurements are also beneficial for covering a whole stomach volume. In this study, with a scan time of 19 s, only three sagittal slices could be acquired with a thickness and interslice gap of 4 and 2 mm, respectively, covering 16 mm in the left-to-right direction. This might not be representative of the whole gastric content, and is a limitation of this feasibility study.

In this study, the intensity thresholding method that we previously applied to *in vitro T_2_*-weighted MRI images (Mayar et al., 2023, 2024) was used to identify low- and high-intensity voxels in *in vivo* images. The low- and high-intensity voxels were attributed to more semi-solid and liquid gastric content, respectively, and were used to quantify their respective volumes over time. It is worth noting that with this approach a threshold value is determined even within 12 min after milk ingestion when milk is still in a liquid state. This stems from the lack of direct mixing between ingested milk and the baseline gastric content, primarily composed of gastric juice. This leads to the formation of a thin layer of baseline gastric content on top of the milk. The baseline gastric content likely comprises gastric juice, based on its higher intensity compared to the ingested milk in the *T_2_*-weighted MRI images. The intensity thresholding will then separate the liquid milk from this high-intensity layer. Therefore, in this study, the obtained semi-solid and liquid volumes were considered from the second timepoint onwards, where coagulation became clearly visible.

A potential limitation of MRI is that it usually requires participants to be in a supine position, which affects the position of the stomach due to gravity. This could lead to a difference in fluid dispersion throughout the stomach, thereby affecting gastric coagulation and emptying (Holwerda et al., 2016). However, relative differences are expected to remain the same (Jones et al., 2006)

### 4.4. Inter-individual variation

This study included both male and female participants for better generalizability. The *MTR*, TGC, and liquid volumes were higher in females as compared to males. However, after accounting for body size, no significant differences remained. This indicates that these differences are related to body size and may not be inherently sex-specific. Previous studies have demonstrated slower GE in females compared to males (Camps, de Graaf, et al., 2018; Gill et al., 1987; Hermansson & Sivertsson, 1996). However, none of these studies included body size as a factor in their statistical analysis. Camps et al. (2018) did find a weak correlation (r= 0.266, p <0.05) between body size and TGC volume, but they concluded that this was not strong enough to explain the effect of sex on TGC volume. Previous research has reported differences in pepsin activity and in gastric pH between males and females (Lindahl et al., 1997), which could also influence gastric coagulation and emptying. Therefore, more research is warranted to better understand the effect of different factors, including body size and (sex) hormones, on gastric digestion.

## 5. Conclusions

We have demonstrated the potential of ^1^H MT MRI as a novel non-invasive method for measuring gastric milk protein coagulation in humans. The *MTR* parameter, which is a marker of coagulum consistency, can be obtained from two rapid scans of less than 20 s each. By combining conventional *T_2_*-weighted anatomical images with MT measurements, the GE of total, semi-solid and liquid gastric content, as well as changes in the coagulum structure, could be assessed *in vivo*.

The *MTR* of LPSM increased during gastric digestion, which reflected milk protein coagulation and was in agreement with previous data from a semi-dynamic *in vitro* model. Interestingly, contrary to the *in vitro* data, no difference was found in the *MTR* between LPSM and HPSM *in vivo*. Moreover, gastric content volumes over time were higher for HPSM compared to LPSM, indicating a slower GE upon heat treatment. These findings demonstrate that the effect of heat treatment on gastric milk protein digestion may be different in humans compared to what has been observed in *in vitro* and animal models. This highlights the importance of conducting *in vivo* studies in humans when investigating effects of heat treatment on gastric milk digestion. Our innovative approach holds promise for investigating the effect of various heat treatments or the presence of lipids on gastric milk protein coagulation and emptying dynamics. Moreover, it opens the way for exploring gastric digestion of a variety of (semi)-solid foods, including yoghurt, cheeses and protein-rich foods with different micro- and macrostructures. Insights from such studies can not only improve our understanding of how food processing, composition, and structure affect gastric digestion, but may also contribute to the improvement of in vitro digestion models.

## Data Availability

All data produced in the present study are available upon reasonable request to the authors

## Author statement

Morwarid Mayar: Conceptualization, Methodology, Formal analysis, Investigation, Data curation, Visualization, Validation, Writing – original draft; Camilla Terenzi; John van Duynhoven and Paul Smeets: Conceptualization, Methodology, Validation, Writing – review & editing, Supervision.

## Acknowledgements

Rosalind Vivia Tansy is gratefully acknowledged for her help with the data collection and analysis. The use of the 3T MRI facility has been made possible by Wageningen University & Research Shared Research Facilities. Paul de Bruin is thanked for his expert support with setting up the MRI measurements.

## References

Ahlborn, N. G., Montoya, C. A., Hodgkinson, S. M., Dave, A., Ye, A., Samuelsson, L. M., Roy, N. C., & McNabb, W. C. (2023). Heat treatment and homogenization of bovine milk loosened gastric curd structure and increased gastric emptying in growing pigs. Food Hydrocolloids, 137, 108380. 10.1016/j.foodhyd.2022.108380

Barbé, F., Ménard, O., Le Gouar, Y., Buffière, C., Famelart, M.-H., Laroche, B., Le Feunteun, S., Dupont, D., & Rémond, D. (2013). The heat treatment and the gelation are strong determinants of the kinetics of milk proteins digestion and of the peripheral availability of amino acids. Food Chemistry, 136(3–4), 1203–1212. 10.1016/j.foodchem.2012.09.022

Bie, C., Liang, Y., Zhang, L., Zhao, Y., Chen, Y., Zhang, X., He, X., & Song, X. (2019). Motion correction of chemical exchange saturation transfer MRI series using robust principal component analysis (RPCA) and PCA. Quantitative Imaging in Medicine and Surgery, 9(10), 1697–1713. 10.21037/qims.2019.09.14

Camps, G., van Eijnatten, E. J., van Lieshout, G. A., Lambers, T. T., & Smeets, P. A. (2021). Gastric Emptying and Intragastric Behavior of Breast Milk and Infant Formula in Lactating Mothers. The Journal of Nutrition, 151(12), 3718–3724. 10.1093/jn/nxab295

Fadda, H. M., Hellström, P. M., & Webb, D.-L. (2022). Intra- and inter-subject variability in gastric pH following a low-fat, low-calorie meal. International Journal of Pharmaceutics, 625, 122069. 10.1016/j.ijpharm.2022.122069

Grimm, M., Koziolek, M., Kühn, J.-P., & Weitschies, W. (2018). Interindividual and intraindividual variability of fasted state gastric fluid volume and gastric emptying of water. European Journal of Pharmaceutics and Biopharmaceutics, 127, 309–317. 10.1016/j.ejpb.2018.03.002

Henkelman, R. M., Stanisz, G. J., & Graham, S. J. (2001). Magnetization transfer in MRI: A review. NMR in Biomedicine, 14(2), 57–64. 10.1002/nbm.683

Holwerda, A., Lenaerts, K., Bierau, J., & van Loon, L. (2016). Body Position Modulates Gastric Emptying and Affects the Post-Prandial Rise in Plasma Amino Acid Concentrations Following Protein Ingestion in Humans. Nutrients, 8(4), 221. 10.3390/nu8040221

Horstman, A. M. H., Ganzevles, R. A., Kudla, U., Kardinaal, A. F. M., van den Borne, J. J. G. C., & Huppertz, T. (2021). Postprandial blood amino acid concentrations in older adults after consumption of dairy products: The role of the dairy matrix. International Dairy Journal, 113, 104890. 10.1016/j.idairyj.2020.104890

Jones, K. L., O’Donovan, D., Horowitz, M., Russo, A., Lei, Y., & Hausken, T. (2006). Effects of Posture on Gastric Emptying, Transpyloric Flow, and Hunger After a Glucose Drink in Healthy Humans. Digestive Diseases and Sciences, 51(8), 1331–1338. 10.1007/s10620-005-9010-3

Lacroix, M., Bon, C., Bos, C., Léonil, J., Benamouzig, R., Luengo, C., Fauquant, J., Tomé, D., & Gaudichon, C. (2008). Ultra High Temperature Treatment, but Not Pasteurization, Affects the Postprandial Kinetics of Milk Proteins in Humans. The Journal of Nutrition, 138(12), 2342–2347. 10.3945/jn.108.096990

Li, S., Pan, Z., Ye, A., Cui, J., Dave, A., & Singh, H. (2022). Structural and rheological properties of the clots formed by ruminant milks during dynamic in vitro gastric digestion: Effects of processing and species. Food Hydrocolloids, 126, 107465. 10.1016/j.foodhyd.2021.107465

Lindahl, A., Ungell, A., Knutson, L., & Lennernäs, H. (1997). Characterization of fluids from the stomach and proximal jejunum in men and women. Pharmaceutical Research, 14(4), 497–502. 10.1023/A:1012107801889

Lu, K., Liu, Z., Jaffey, D., Wo, J. M., Mosier, K. M., Cao, J., Wang, X., & Powley, T. L. (2022). Automatic assessment of human gastric motility and emptying from dynamic 3D magnetic resonance imaging. Neurogastroenterology & Motility, 34(1). 10.1111/nmo.14239

Mayar, M., de Vries, M., Smeets, P., van Duynhoven, J., & Terenzi, C. (2024). MRI assessment of pH and coagulation during semi-dynamic in vitro gastric digestion of milk proteins. Food Hydrocolloids, 109866. 10.1016/j.foodhyd.2024.109866

Mayar, M., Miltenburg, J. L., Hettinga, K., Smeets, P. A. M., van Duynhoven, J. P. M., & Terenzi, C. (2022). Non-invasive monitoring of in vitro gastric milk protein digestion kinetics by 1H NMR magnetization transfer. Food Chemistry, 383(February), 132545. 10.1016/j.foodchem.2022.132545

Mayar, M., Smeets, P., Duynhoven, J., & Terenzi, C. (2023). In vitro 1H MT and CEST MRI mapping of gastro-intestinal milk protein breakdown. Food Structure, 36, 100314–100324.

Milan, A. M., Barnett, M. P., McNabb, W. C., Roy, N. C., Coutinho, S., Hoad, C. L., Marciani, L., Nivins, S., Sharif, H., Calder, S., Du, P., Gharibans, A. A., O’Grady, G., Fraser, K., Bernstein, D., Rosanowski, S. M., Sharma, P., Shrestha, A., & Mithen, R. F. (2024). The impact of heat treatment of bovine milk on gastric emptying and nutrient appearance in peripheral circulation in healthy females: a randomized controlled trial comparing pasteurized and ultra-high temperature milk. The American Journal of Clinical Nutrition. 10.1016/j.ajcnut.2024.03.002

Mulet-Cabero, A. I., Mackie, A. R., Wilde, P. J., Fenelon, M. A., & Brodkorb, A. (2019). Structural mechanism and kinetics of in vitro gastric digestion are affected by process-induced changes in bovine milk. Food Hydrocolloids, 86, 172–183. 10.1016/j.foodhyd.2018.03.035

Roelofs, J. J. M., Camps, G., Leenders, L. M., Marciani, L., Spiller, R. C., Eijnatten, E. J. M. van, Alyami, J., Deng, R., Freitas, D., Grimm, M., Karhunen, L. J., Krishnasamy, S., Feunteun, S. Le, Lobo, D. N., Mackie, A. R., Mayar, M., Weitschies, W., & Smeets, P. A. M. (2024). Intra- and interindividual variability in fasted gastric content volume. MedRxiv, 2024.03.12.24304085. 10.1101/2024.03.12.24304085

Semple, J. I., Newton, J. L., Westley, B. R., & May, F. E. B. (2001). Dramatic diurnal variation in the concentration of the human trefoil peptide TFF2 in gastric juice. Gut, 48(5), 648–655. 10.1136/gut.48.5.648

van Eijnatten, E. J. M., Camps, G., Guerville, M., Fogliano, V., Hettinga, K., & Smeets, P. A. M. (2023). Milk coagulation and gastric emptying in women experiencing gastrointestinal symptoms after ingestion of cow’s milk. Neurogastroenterology & Motility. 10.1111/nmo.14696

van Zijl, P. C. M., Lam, W. W., Xu, J., Knutsson, L., & Stanisz, G. J. (2018). Magnetization Transfer Contrast and Chemical Exchange Saturation Transfer MRI. Features and analysis of the field-dependent saturation spectrum. NeuroImage, 168, 222–241. 10.1016/j.neuroimage.2017.04.045

Ye, A., Cui, J., Dalgleish, D., & Singh, H. (2016). The formation and breakdown of structured clots from whole milk during gastric digestion. Food & Function, 7(10), 4259–4266. 10.1039/C6FO00228E

Ye, A., Roy, D., & Singh, H. (2019). Structural changes to milk protein products during gastrointestinal digestion. In Milk Proteins: From Expression to Food (3rd ed.). Elsevier Inc. 10.1016/B978-0-12-815251-5.00019-0

Zhou, Y., Bie, C., van Zijl, P. C. M., & Yadav, N. N. (2023). The relayed nuclear Overhauser effect in magnetization transfer and chemical exchange saturation transfer MRI. NMR in Biomedicine, 36(6). 10.1002/nbm.4778

