## Supplementary Information for "Magnetization Transfer MRI of intragastric milk digestion: a feasibility study in humans"

For

1. Flow diagram of the study

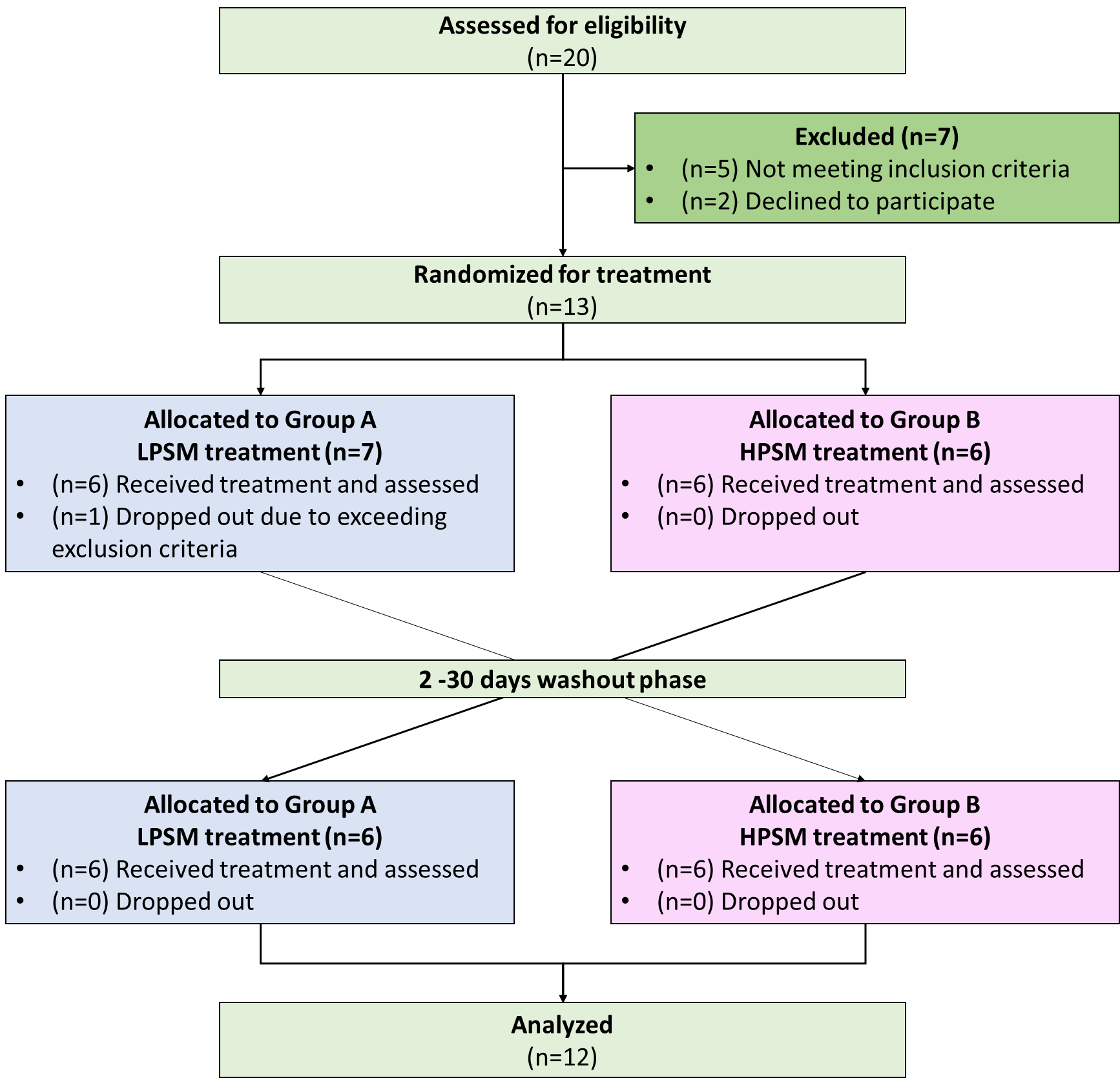

***Figure S1.*** *Flow diagram for inclusion, treatment allocation and analysis.*

2. Sample size estimation

This is the first study in which MT MRI will be applied to study *in vivo* gastric protein digestion, and hence, the expected differences and population variance are unknown. Therefore, we use our *in vitro* findings of the evolution in semi-solid MTR during semi-dynamic *in vitro* digestion to estimate the expected difference and variance. The standard deviation (SD) *in vitro* was 0.0032 and, assuming that the SD *in vivo* will be at least 30 times higher due to physiological noise and movement-related distortions, we estimate a SD of 0.096. A repeated measures ANOVA will be used to determine if there is significant effect of time and treatment on the MTR. With a power of 90%, and a significance level of 0.05, a total of at least 9 or 12 complete datasets is required for the time and treatment effect, respectively (calculated using: https://glimmpse.samplesizeshop.org/). Therefore, a total of 12 participants will be recruited to be able to evaluate both the time and treatment effect. Participants will be replaced in case of dropouts up until a maximum of 14 participants.

3. Image registration and intensity thresholding

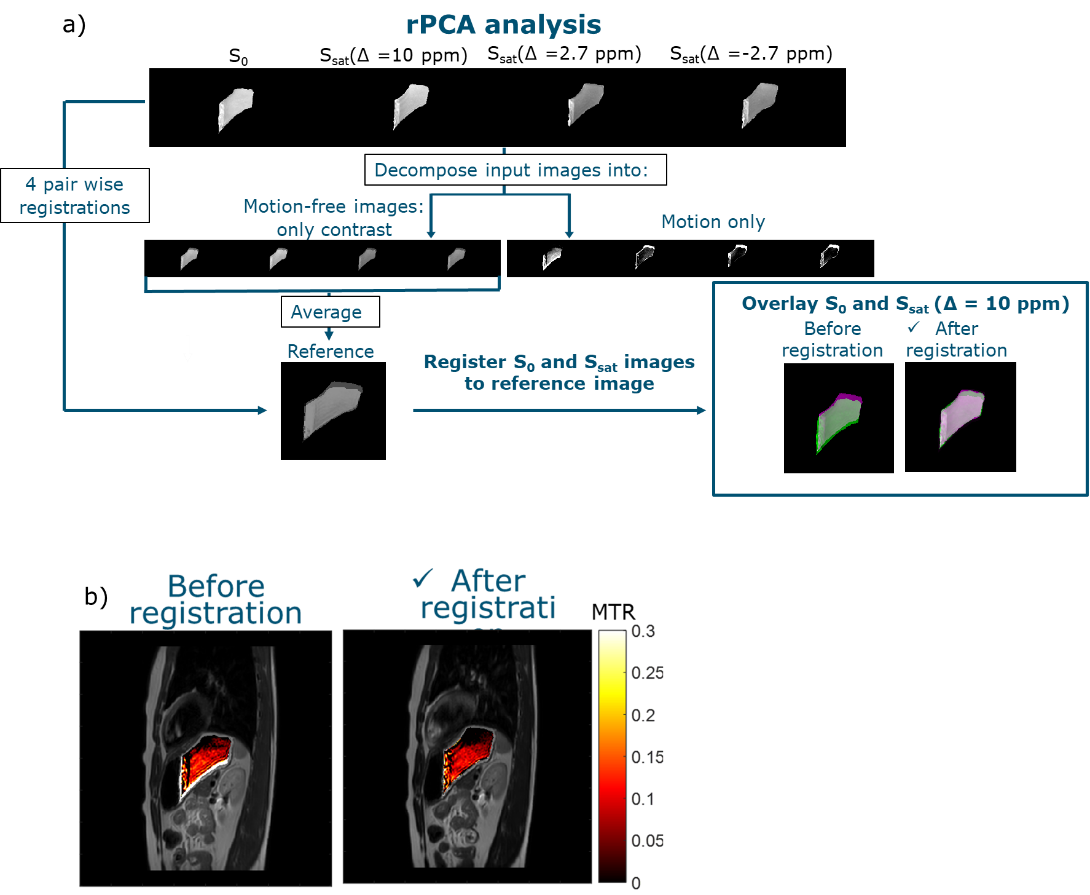

***Figure S2****. (a) Schematic overview of the image registration process of the masked gastric content images, consisting of decomposing the MT images into a low-rank and sparse component, containing the motion-free contrast and motion information, respectively. The average of the low-rank images is used as a reference image for alignment of the original motion-corrupted images, resulting in well-aligned S_0_ and S_sat_ images. (b) MTR maps of the gastric content overlayed on the T_2_-weighted MRI image at t = 5 min after ingestion of 300 g of milk before (left) and after (right) image registration.*

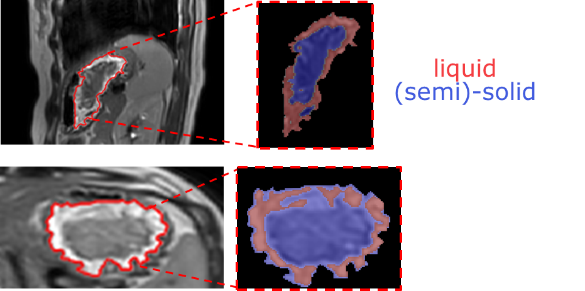

***Figure S3****.Liquid and semi-solid masks obtained by intensity thresholding of the sagittal (top) and axial (bottom) T_2_-weighted MRI images.*

4. MTR data

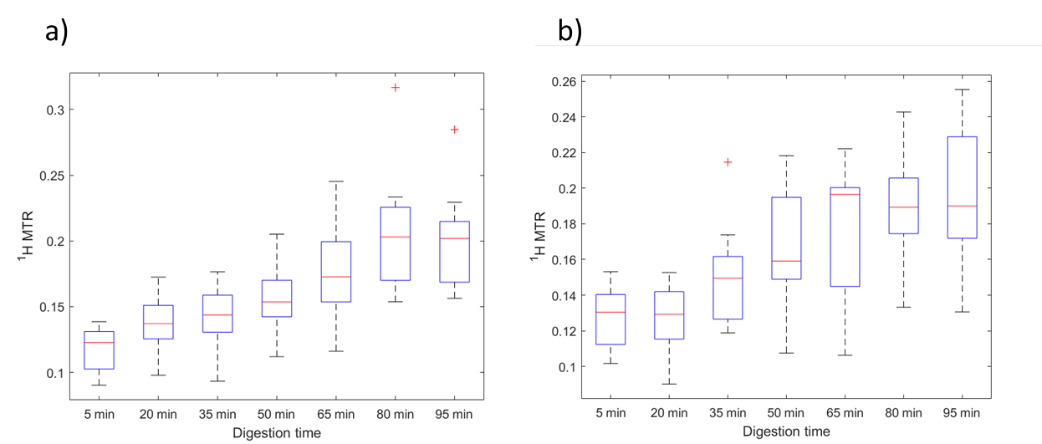

***Figure S4****. Boxplots of the MTR over time for (a) LPSM and (b) HPSM. Outliers are shown are marked with a red symbol (+).*

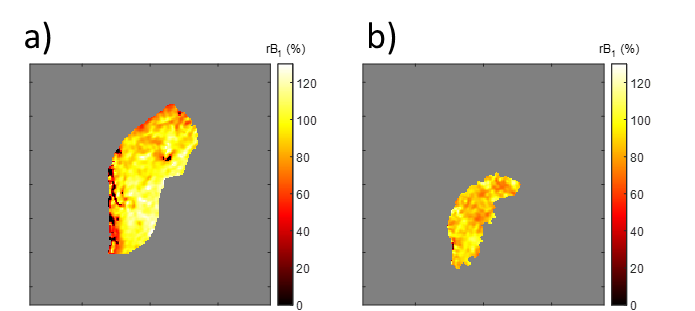

***Figure S5****. rB₁ maps of the gastric content at (a) t = 5 min and (b) t = 95 min after the ingestion of 300 g HPSM are depicted for the same participant as shown in Fig. 5.4 in the main text. These maps illustrate a decrease in rB₁ in the gastric content from t = 5 to 95 min. Specifically, for the voxels used to calculate the MTR, this decrease was from 100±15% at t = 5 min to 87±9% at t = 95 min.*

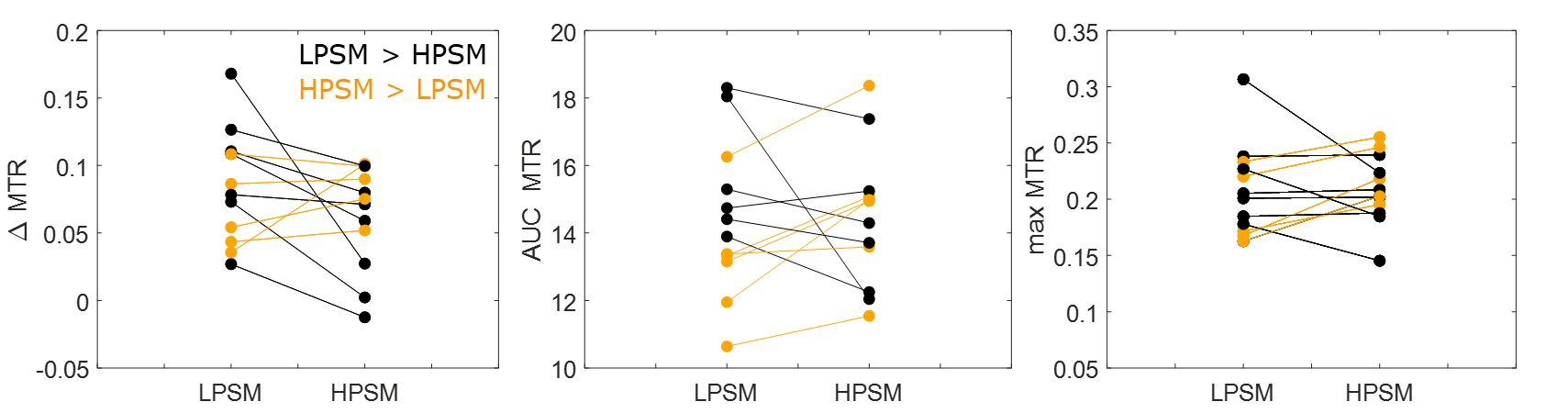

***Figure S6.*** *(a) ΔMTR between T5-95 min, (b) AUC MTR and (c) max MTR per participant for LPSM and HPSM.*

5. Effect of sex on digestion outcomes

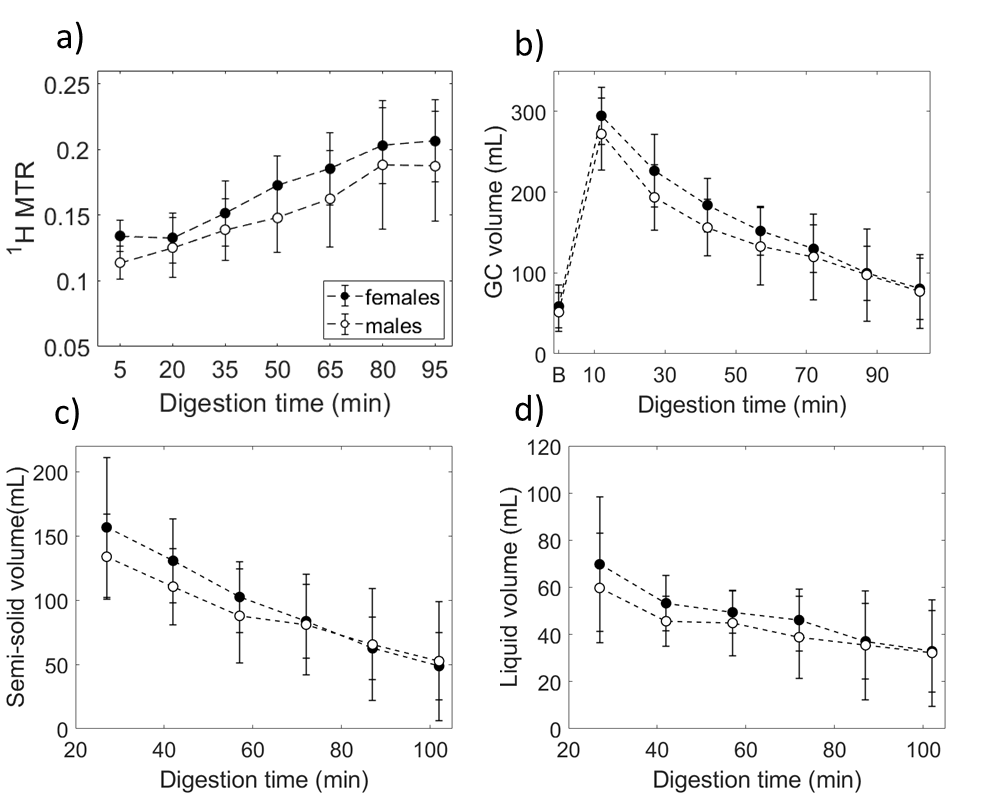

***Figure S7.*** *(a) MTR, (b) GC volume, (c) semi-solid volume and (d) liquid volume for female and male participants. The data of LPSM and HPSM were grouped per sex. Data are show as mean±SD.*

6. Appetite and well-being ratings

| 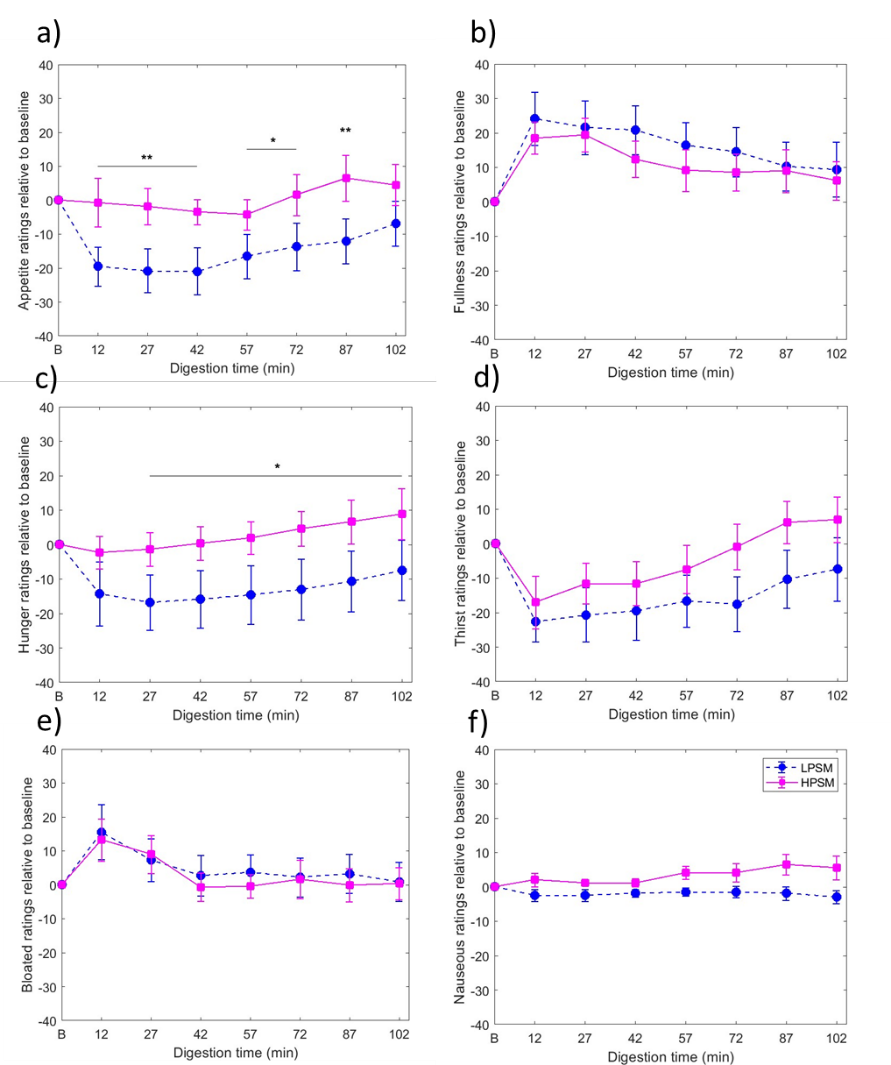 |
| --- |

***Figure S8.*** *Changes relative to baseline in ratings (0-100 points) of the subjective feeling of hunger, fullness, appetite, thirst, bloating, and nausea over time. All values are presented as mean ± SEM (n = 12). *p<0.05 and **p<0.01.*

***Table S1.*** *Pearson correlation analysis between the baseline MTR values and the* ${MTR}_{max}$

*and* $AUC MTR$*.*

|  | ${MTR}_{max}$ | $AUC MTR$ |
| --- | --- | --- |
| LPSM | r = 0.09; p =0.80 | r = 0.095; p = 0.79 |
| HPSM | r = 0.2; p =0.56 | r = 0.33; p = 0.33 |
